# Quantifying the preventive effect of wearing face masks

**DOI:** 10.1101/2021.02.09.21251448

**Authors:** Tom Britton

## Abstract

An important task in combating the ongoing covid-19 effect lies in estimating the effect of different preventive measures. Here we focus on the preventive effect of enforcing the use of face masks. Several publications study this effect, however often using different measures such as: the relative attack rate in case-control studies, the effect on incidence growth/decline in a specific time-frame, the effect on the number of infected in a given time-frame. These measures all depend on community-specific features and are hence not easily transferred to other community settings. We argue that a more universal measure is the relative reduction in the reproduction number, which we call the *face mask effect, E*_*FM*_. It is shown how to convert the other measures to *E*_*FM*_. We also apply the methodology on three publications using different other measures (two of them resulting in two different estimates each, and all five estimates of *E*_*FM*_ lie between 20-40%, suggesting that mandatory face masks reduce the reproduction number by 20-40% as compared to no individuals wearing face masks.

## >1 Introduction

A main motivation for modelling and statistical analyses of infectious disease outbreaks is to understand and to estimate the effect of introducing different preventive measures. Prior to Covid-19, the type of preventive measure which by far had received most attention on how to quantify and estimate its different effects is vaccination. There are e.g. recipies for estimating the vaccine efficacy with respect to: susceptibility (*V E*_*S*_), infectivity (*V E*_*I*_), disease/symptoms *V E*_*D*_, overall *V E*_*all*_, and more, under various settings (e.g. [1, 7]).

Here our focus lies on quantifying the effect of wearing face mask in the ongoing Covid-19 pandemic. More specifically, we aim at quantifying the effect of making face mask mandatory in a community when, compared to no individuals wearing face masks. Several studies from different regions estimate the effect of face mask regulations, but most of them use efficacy measures which are specific the region of interest and which hence are hard to transfer to other settings.

The purpose of the present paper is to define a face mask effect being directly linked to the reduced risk of getting infected when wearing a face mask *and* to the reduced risk of infecting others when wearing a face mask, thus making it more easily transferable to other community settings. Using modelling results from epidemic theory we then describe how to derive estimates of this effect from other community specific efficacy measures. We apply the methodology on three studies which come from different regions and which also use different types of data to quantify the preventive effect of face masks.

## 2 The face mask effect *E*_*FM*_

When it comes to effects of wearing face masks the two most direct effects are the efficacy with respect to susceptibility *e*_*S*_, and the efficacy with respect to infectivity *e*_*I*_. By *e*_*S*_ we mean the reduced risk of infection in a contact between an infective without face mask and a susceptible wearing a face mask as compared to the susceptible not wearing a face mask, so *e*_*S*_ = 1*−P* (*i*_*No*_ *→ s*_*FM*_)*/P* (*i*_*No*_ *→ s*_*No*_) where “→” means an infection occurs in a contact and the index on *i* and *s* reflect whether the susceptible/infective wears a face mask or not. Similarly, by *e*_*I*_ we mean the reduced risk of infection in a contact with a susceptible not wearing a face mask when the infective wears a face mask as compared to when the infective does not wear a face mask, so *e*_*I*_ = 1 *−P* (*i*_*FM*_ *→ s*_*No*_)*/P* (*i*_*No*_ *→ s*_*No*_).

Further, we make the natural assumption that these two effects act multiplicatively, so that the reduced risk of infection if both individuals wear face mask as compared to when neither of them do equals 1 *−P* (*i*_*FM*_ *→ s*_*FM*_)*/P* (*i*_*No*_ *→ s*_*No*_) = 1 − (1 *−e*_*S*_)(1*−e*_*I*_). Since all transmission probabilities are reduced by this factor when all individuals go from not wearing a face mask to all wearing face masks, it follows that this effect will also be the relative reduction in the reproduction number *R* in a community which goes from no one wearing a face mask to all individuals wearing a face mask. We call this the face mask effect *E*_*FM*_ :

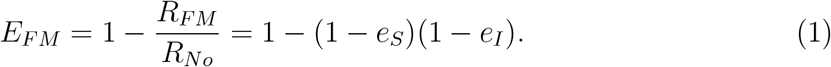

It is important to note that this effect is independent of the underlying reproduction number *R*_*No*_ which typically depends on which community is studied, what other preventive measures are currently in force, and how much immunity the community currently has.

## >3 The modelling perspective

We now derive the effect of introducing face mask regulation on other quantities of an ongoing outbreak, using theory for epidemic models (e.g. [5]). We do this under the assumption that other aspects of the epidemic (e.g. preventive measures, immunity, seasonal effects) remain unchanged, thus implicitly assuming a study period of weeks rather than several months.

### >3.1 Effect on the exponential growth/decline rate

For an ongoing epidemic which currently has *I*(*t*) infectious individuals and current reproduction number *R* (be it with or without mandatory face masks), is known to progress exponentially:

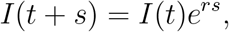

where *r* is known as the Malthusian parameter. This exponential growth (or decay) depends on the reproduction number *R* but also on the generation time distribution *g*(*s*) (which describes the random time between getting infected and infecting others). The relation between *r* and *R* and *g*(*s*) is given by the Euler-Lotka equation (e.g. [3]), and if we make the pragmatic and common assumption that the generation time follows a Gamma distribution with mean *µ* and standard deviation *σ*, then the Euler-Lotka equation is given by

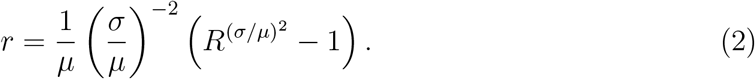

The equation becomes even simpler if we introduce *c* = *σ/µ*, the coefficient of variation of the generation time distribution: 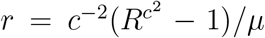 (*σ* = 0 is not allowed since infections would only happen at prespecified time points). From this equation it is seen that *r <* 0 (decline) if *R <* 1 and *r >* 1 (growth) if *R >* 1.

The exponential growth/decline rate without face mask, *r*_*No*_ should hence be computed with *R*_*No*_ and the exponential growth/decline rate with mandatory face masks, *r*_*F M*_, should be computed with *R*_*F M*_. If we compare the growth rate with and without mandatory face masks it is seen that this relation will not only depend on the face mask effect *E*_*F M*_ = 1 *− R*_*F M*_ */R*_*No*_, but it will also depend on the value of the underlying reproduction number *R*_*No*_ (and also on *µ* and *σ*). As a consequence, the effect of face mask on the exponential rate *r* will not be the same in different communities, nor for the same community if comparing different time points of introducing face mask regulation if other preventive measures may have changed. When estimating *E*_*F M*_ we will want to express *R* in terms of the exponential growth rate. For this we invert Equation 2:

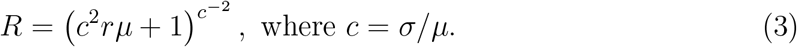

### 3.2 Effect on change of incidence during a given time frame

The incidence *i*(*t*) is defined as the number of new infections per day. This incidence is more or less always assumed to proportional to the current number of infectious individuals *I*(*t*) (the more infectious people the higher infection pressure). As a consequence the ratio of the incidence at time *t* + *d* and the incidence at *t* will equal

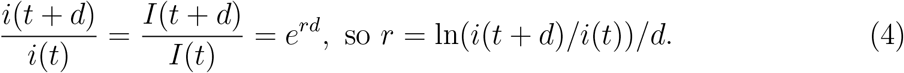

If the time frame (*t, t* + *d*) has no face mask regulation then *r*_*No*_ should be used, whereas *r*_*F M*_ should be used if face masks are mandatory during this period. Just like for the face mask effect on the exponential rate *r*, the face mask effect on the change of incidence-ratio will not only depend on *E*_*F M*_ but also on the underlying reproduction number *R*_*No*_. It follows that different communities typically will see different effect on incidence ratios, even when the same duration *d* of time frames are used.

### >3.3 Effect on number of infections during a given time frame

We now describe how the number of intections within a given time frame is affected by face mask regulation as compared to no one wearing face masks. If *N* (*t*) denotes the number of infected individuals up to time *t* we are hence interested in how *N* (*t* +*d*) *−N* (*t*) changes dependning on whether face masks are mandatory or not. This number depends on the exponential growth rate, but also on *I*(*t*), the starting number of infectious individuals. As before we assume that the incidence *i*(*t*) = *N* ^*‘*^(*t*) is proportinal to *I*(*t*): *N* ^*‘*^(*t*) = *λI*(*t*) where *λ* is the mean infection rate for infectives. From before we know that *I*(*t*) grows exponentially so that *I*(*t* +*s*) = *I*(*t*)*e*^*rs*^. As a consequence, we get the following expression for the (expected) number of infections during the studied time frame:

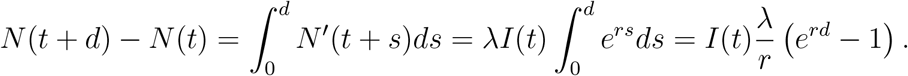

Putting observable quantities on one side we get 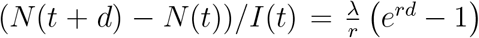. This equation contains a new “parameter” *λ*. In order to obtain a numerical value for *λ* we make an additional approximation by relating *λ* to the mean duration of the infectious period under the assumption that the underlying epidemic model is the Markovian SIR epidemic model (which assumes no latent period and an exponential infectious period). For this case *r* = *λ − γ* where 1*/γ* equals the mean generation time earlier denoted *µ*. We hence have that *λ/r* = 1 + 1*/*(*µr*), and we get

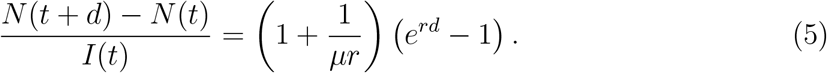

If the study period occurs when the community is not wearing face masks it should be evaluated with *r*_*No*_, and using *r*_*F M*_ if face masks are mandatory. Just like with the incidence ratio, the face mask effect on number of infected not only depends on the starting number of infectives and the duration of the study period, but also on other community settings. This face mask effect is hence not transferable to other community settings either.

## >4 Estimation of *E*_*FM*_ from different empirical data sources

We now make use of the modelling results in the previous section to obtain estimates of the face mask effect *E*_*F M*_ from empirical studies estimating other measures of the effect of wearing face mask.

### >4.1 Estimating *E*_*FM*_ from a case-control studiy

A Danish study [4] was based on a case control study in which approximately 3000 randomly selected individuals were instructed to follow all recommendations *and* to wear a face mask, and another 3000 individuals were instructed to follow all recommendation but to *not* wear a facemask. All indivduals were susceptible at the start of the study period in April 3, and after 2 months they were tested for infection. There were some drop-outs during the study, so the final result was that 42 out of 2392 assigned to wear face masks were infected, and in the other group 53 out of 2470 got infected. The (small) risk of getting infected during the study period was hence 1.8% (42/2390) for individuals wearing a face mask, and 2.1% (53/2470) for the control group. The reduced risk of getting infected in case of wearing a face mask as compared to not wearing a face mask is hence estimated to equal 18% (=1-0.018/0.021). The two groups were by no means separated in the community, so the reduced risk is attributed to reduction in susceptibility – the potential reduction in infectivity (if infected) cannot be estimated from such a study.

The study has two important advantages. The first is that individual were randomly distributed to the two groups, thus making the risk for confounding effects negligible. The other advantage (which is also a disadvantage!) was that the risk of getting infected was quite small. As a consequence, since the vast majority did not get infected it is reasonable to interpret the reduction by 18% as a reduction effect *per contact*, so that *e*_*S*_ can be estimated *ê*_*S*_ = 0.18. If a bigger fraction would have gotten infected, individuals would have been exposed to infections many times, and there would be no reason to expect the fraction infected among individuals wearing face mask to be *e*_*S*_ lower than among individuals not wear a face mask.

Because the total number infected in both arms were small the estimated effect of *ê*_*s*_ = 18% has a wide confidence interval spanning from −23% up to +46% (a negative effect would imply that wearing a face mask *increases* the risk of getting infected), and the study does hence not show that the reduction in susceptibility is statistically significant.

Still, the point estimate *ê*_*S*_ = 0.18 indicates a protective effect for the risk of getting infected, but the study gives no information on the reduction in infectivity *e*_*I*_. Experimental studies (e.g. [8]) indicate that *e*_*I*_ *> e*_*S*_, so a conservative estimate of the overall preventive effect of wearing face masks would be to assume *e*_*I*_ = *e*_*S*_ which gives the estimated face mask effect

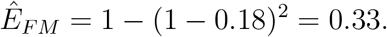

This estimate is of course equipped with high uncertainty, but the point estimate at least suggests that the reproduction number is reduced by approximately 33%. If we instead would assume that the reduction in infectivity is 50% higher than that of susceptibility, then the estimate of *E*_*F M*_ would be 1 − (1 − 0.18)(1 − 0.27) = 0.40, an even bigger effect.

### >4.2 Estimating *E*_*FM*_ from incidence change in a retrospective study

A more common type of study when estimating the face mask effect are retrospecitive studies. A potential problem with these type of studies is of course that even if attempts are made to remove potential biases, there is always a risk that the communities compared, or the observed time periods differ also in other aspects than use of face masks.

One such study [10] compared different counties in Kansas, USA, some which had enforced face mask regulations and other counties which did not. The regulations were put in force on July 5, 2020, and it was studied how the incidence grew before the change and after the change, and these changes in growth rate were compared to changes in growth rate for the same periods in counties which did not enforce wearing of face masks. We postpone estimation from comparing different regions to the next subsection (for another study) and here focus on the change of incidence over time before and after the introduction of face mask regulation.

The study showed that in counties later having mandatory face masks the average incidence in early June was 3, in early July it had risen to 17 (after which regulations were put in place), and in mid-August the incidence rate was 16, so the incidence rate stopped growing after face mask regulation were put in place.

The study reports that incidence increased by 0.25 cases per 100 000 individuals per day prior to face mask regulation whereas incidence *decreased* by 0.08 cases per 100 000 individuals per day after face mask became mandatory. Eventhough these numbers are completely true they carry very little information on what might be the effect in a different community. For this reason we now estimate the face mask effect *E*_*F M*_ from the same data, using the methods described in the modelling section.

Since communities without face mask regulations had no drop in exponential growth rate (in fact a small increase), we attribute the change in incidence in communities adopting face mask regulation entirely to this effect (as in [10]).

The incidence was reported to equal 3.0 cases per 100 000 individuals on June 3 (weekly average with June 3 as midpoint) and incidence 17.0 cases per 100 000 individuals on July 5, 32 days later. Using (4) with *d* = 32 we obtain an estimate of the exponential growth prior to face mask regulation as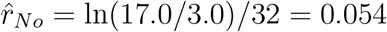. In order to estimate *R*_*No*_ (and eventually *E*_*F M*_) using (2) we need to make some assumptions about the generation time and we choose *µ* = 6.5 days and *σ* = 4 days as in [6]. By inverting (2) we obtain the estimate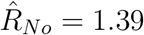.

The growth rate after regulations were more or less nill, or in fact a very small decline (from 17 to 16 in 45 days). This suggests a reproduction number very close to one; using the same methodology we get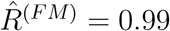. The estimate of the overall effect of wearing face mask from the Kansas study is hence

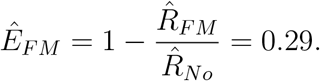

The Kansas study [10] hence suggests that the effect of enforcing face masks to be worn has the overall preventive effect *Ê*_*F M*_ = 29%. This estimate is equipped with less uncertainty than the Danish study but may on the other hand have confounding factors resulting in a biased estimate. For instance, a difference between counties imposing face mask regulations early and counties who did not was that the former had experienced a quicker epidemic growth in June. Most likely this partly explains their willingness to enforce face masks, but there is of course also a possibility that individuals in these communities became more precautious compared to individuals in communities which experienced lower growth rates. If this was the case, the estimated effect would only partly be attributed to face masks.

### >4.3 Estimating *E*_*FM*_ from change of number of infections in a retrospective study

Another recent retrospective study [9] analyses the effect that an early face mask regulation had in the German city of Jena, as compared to a “synthetic control group” defined with the aim to minimize other confounding effects. One of the main results was that the number of reported cases (per 100 000 individuals) during a 20 day time window dropped by 75% when comparing Jena with the synthetic control group. More precisely, the cumulative number of reported cases in Jena on April 6 when face mask regulation was introduced was 42 (per 100 000 individuals), and 158 reported cases on April 26. The corresponding numbers for the synthetic control group were 142 and 205. The number of infections in the time window was hence 16.0 in Jena and 63.0 in the synthetic control group (per 100 000 individuals), a drop of 75% in Jena compared to the synthetic control group. In order to estimate the epidemic growth we also needed to know the number of infectious people at the start of the study period (it it is low suggest a quick growth and the opposite if the number of infectious would be high). This inforation is of course harder to obtain, but it we assume the infectious period is approximately 4 days (which if preceded by a 3 day latent period is in agreement with [6]) then this number can be estimated from the reported number of cases prior to April 6. These numbers are close to identical for Jena and the control group and since the number of reported cases is very clsoe to 4 per day (per 100 000 individuals) we estimate the number of infectious individuals on April 6 to be 16 for both Jena and the synthetic control group.

Using Equation 5 we can now estimate *r*_*F M*_ for Jena and *r*_*No*_ for the synthetic control group. For Jena the left hand side equals 16*/*16 and the right hand side has *r* as unknown and *µ* = 6.5 and *d* = 20 (the length of the time window) giving the estimate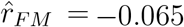, and for the synthetic control group the right hand side is the same but the left hand side is 63*/*16, giving the estimate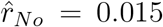. Using 3 with *µ* = 6.5 and *c* = 4*/*6.5 = 0.615 we get 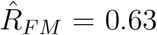 for Jena and 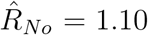 for the synthetic group, thus resulting in an estimate of *E*_*F M*_ of 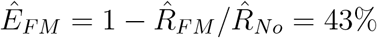.

This estimate of the face mask effect *E*_*F M*_ is notably higher than the other two estimates of *E*_*F M*_ which both are close to 30%. However, Mitze et al. [9] argue that the decrease by in number of infections by 75% might be an over-estimate and when taking other aspects into account (including effect of face mask regulations in other regions later on) suggest a better estimate of the reduction in number of infections attributed to face mask regulation be 45%, which would correspond to 34.6 infections (per 100 000 inhabitants) rather than 16. If instead this number was used in the estimation procesure, the face mask effect would be estimated to *Ê*_*F M*_ = 21%.

## >5 Conclusions and Discussion

Using results from mathematical theory for epidemic models we relate different observable quantities with the reproduction number *R*. By inverting these relations it is possible to obtain estimates of the reproduction number, with and without face mask, from empirical studies investigating such quantities. Three studies were used to obtain estimates of the face mask effect *E*_*F M*_ = 1 *− R*_*F M*_ */R*_*No*_, where *R*_*F M*_ is the reproduction number *with* enforced face masks, and *R*_*No*_ is the reproduction number when individuals are not using face masks. In Table 1 we summarize our results. The Danish study [4] results in two estimates of *E*_*F M*_ depending on if the reduced risk of infecting others when wearing face mask, *e*_*I*_, is equal or 50% higher than the reduced risk of getting infected by wearing a face mask *e*_*S*_. Also the German study [9] has two estimates of *E*_*F M*_ depending on if only Gena is used for estimating *E*_*F M*_ or if also estimates from other cities are used (as suggested in [9]).

**Table 1:** Estimates of the face mask effect *E*_*F M*_ from different empirical studies

| Reference/country | Type of reported data | Submodel | Estimated $E_{FM}$ |
| --- | --- | --- | --- |
| [4] Denmark | Relative attack rate | $e_I = e_S$ | 33% |
| [4] Denmark | Relative attack rate | $e_I = 1.5e_S$ | 40% |
| [10] USA | Change of incidence |  | 29% |
| [9] Germany | Change in reported cases | Jena only | 43% |
| [9] Germany | Change in reported cases | Combined estimates | 21% |

As expected, the different estimates show some variation. Still, they suggest that the effect of introducing face mask regulation reduces the reproduction number by 20-40%, a rather big reduction for one single preventive measure. Even in the lower end of this interval, a community not using face masks and currently having *R*_*No*_ = 1.2 (with a doubling time of daily cases being less than a month) would change its reproduction number to *R*_*F M*_ = 0.96 *<* 1 if face masks were made mandatory, hence experiencing a slow decline in incidence instead. If instead *EFM* = 40% the new reproduction would be *R*_*F M*_ = 0.72 and transmission would drop quickly in the community.

The estimates above are estimates when going from no face masks to all individuals wearing face masks. If there in some community are already a fraction *f*_*B*_ wearing face masks before the regulation is put in place, and the recommendation/regulation results in that a higher fraction *f*_*A*_ then wear face masks. Then an estimate of this effect would be (*f*_*A*_ *− f*_*B*_)*E*_*F M*_, so if for example a recommendation results in that the fraction wearing face masks increases from 20% to 70%, then the effect would be 0.5 *∗ E*_*F M*_.

We have found only one other study which estimates the same efficacy measure *E*_*F M*_ as suggested here and this is [2]. This is a type of meta analysis study based on data from 190 countries. Their estimate equals *Ê*_*F M*_ = 15.1%, so slightly lower than our observed interval 20%-40%, but their estimate equals 33% before adjusting for confounding factors, something which is very hard when analysing many countries jointly. The observed difference should not be interpreted as a contradiction, but rather that it is hard to estimate *E*_*F M*_ from empirical studies whatever method is used since confounding factors may always effec the estimates.

Even if the suggested measure *E*_*F M*_ is claimed to be “universal” this is not entirely true. For example, if people adhere strictly to the face mask regulation, also outdoor and when at home in one community, but in another community people do not wear face masks in their own home or when outdoors, then the face mask effect will of course be higher in the former community. The individual face mask efficacy also depends on whether or not the face mask is worn properly, and if it is removed and put on in a safe way, and clearly the type of face mask and how well it fits the user will also affect the individual efficacy. However, unless different communities are systematically different in these aspects, the suggested measure *E*_*F M*_ is a population average effect which should not vary greatly between communities.

## Data Availability

No data is used except already published data from other scientific papers.

## Acknowledgements

The author is grateful for financial support from NordForsk, grant 105572.

## References

[1] Becker N.G., Britton T. and O’Neill P.D. (2003): Estimating vaccine effects on transmission of infection from household outbreak data. Biometrics. 59: 467–475.

[2] Bo, Y., Guo, C., Lin, C. et al. (2020). Effectiveness of non-pharmaceutical interventions on COVID-19 transmission in 190 countries from 23 January to 13 April 2020. Int. J. Inf. Dis. 102: 247–253.

[3] Britton, T. and Scalia Tomba G. (2019). Estimation in emerging epidemics: biases and remedies. Journal Royal Society: Interface. 16:20180670, https://doi.org/10.1098/rsif.2018.0670.

[4] Bundgaard, H., Skov Bundgaard, J., Raaschou-Pedersen, D.E.T, et al. (2020). Effectiveness of Adding a Mask Recommendation to Other Public Health Measures to Prevent SARS-CoV-2 Infection in Danish Mask Wearers. An. Int. Med. https://doi.org/10.7326/M20-6817

[5] O. Diekmann, H. Heesterbeek, T. Britton, Mathematical tools for understanding infectious disease dynamics, (Princeton University Press, 2013).

[6] Flaxman, S., Mishra, S., Gandy, A. et al. (2020). Estimating the effects of nonpharmaceutical interventions on covid-19 in Europe. Nature, 584: 257261.

[7] Halloran, M.E., Longini, I.M., and CJ Struchiner, C.J. (2010). Design and analysis of vaccine studies. Springer.

[8] Kähler, C.J. and Hain, R. (2020). Fundamental protective mechanisms of face masks against droplet infections. J. Aerosol Sci. 148: 105617.

[9] Mitze, T., Kosfeld, R., Rode, J. and Wlde, K. (2020). Face masks considerably reduce COVID-19 cases in Germany. P.N.A.S., 117:51, 3229332301.

[10] Van Dyke, M.E., Rogers, T.M., Pevzner, E., et al. (2020). Trends in county-level COVID-19 incidence in counties with and without a mask mandateKansas, June 1August 23, 2020. Morbidity and Mortality Weekly Report (DCD), 69:47.

